# Value of serum albumin, age, serum creatinine, and left ventricular ejection fraction for the assessment of 4-year mortality risk in patients with acute myocardial infarction: parsimonious and better performed

**DOI:** 10.1101/2022.08.07.22278501

**Authors:** Zheng-Yang Ge, Yang He, Ting-Bo Jiang, Jian-Ying Tao, Yong-Ming He

## Abstract

**Aims:** Mortality from acute myocardial infarction (AMI) remains substantial. The current study is aimed at developing a novel simple and easy-to-use risk score for AMI.

**Methods:** The CatLet extended validation trial (ChiCTR2000033730) and the CatLet validation trial (ChiCTR-POC-17013536), both being registered with chictr.org, served as the derivation and validation datasets, respectively. The derivation dataset included 1018 patients, and the validation dataset included 308 ones. They all suffered from AMI and underwent percutaneous intervention (PCI). The 4-year follow-up was 97% completed for the derivation dataset, and 100% completed for the validation dataset. The endpoint was all-cause death. Lasso regression analysis was used for covariate selection and coefficient estimation.

**Results:** Of 26 candidate predictor variables, four strongest predictors for 4-year mortality were included in the BACEF score (serum albumin, age, serum creatinine, and LVEF). This score was well calibrated and yielded an AUC (95%CI) statistics of 0.84(0.80-0.87) in internal validation, 0.89(0.83-0.95) in internal-external (temporal) validation, and 0.83(0.77-0.89) in external validation. Notably, it outperformed the ACEF, ACEFII, GRACE risk scores (0.83(0.77-0.89) vs 0.81(0.75-0.88); 0.83(0.77-0.89) vs 0.79(0.73-0.86); and 0.83(0.77-0.89) vs 0.79(0.72-0.86), respectively).

**Conclusions:** A simple risk score for 4-year mortality risk stratification was developed, extensively validated, and calibrated in patients with AMI. This novel BACEF score outperformed the ACEF, ACEFII, and GRACE scores and may be a useful risk stratification tool for patients with AMI.

**One-sentence summary:** A novel simple risk score with an acronym of BACEF, including only four risk factors of serum albumin, age, serum creatinine, and LVEF, has been developed, and outperformed the ACEF, ACEFII, and GRACE scores in terms of four-year mortality prediction for patients with acute myocardial infarction.

**Take-home figure:** 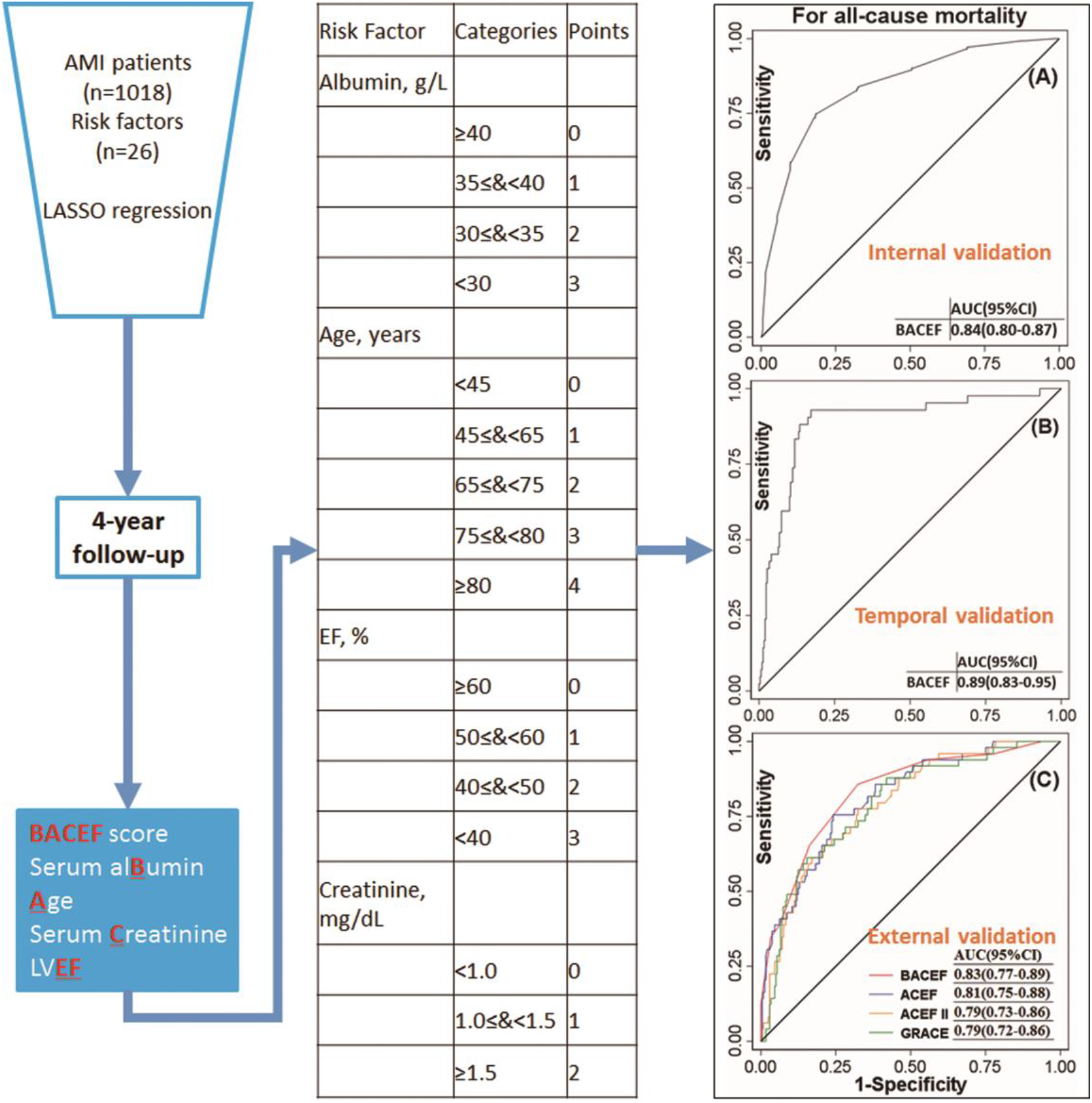

## Introduction

Several risk scores have been introduced and are currently in use for acute coronary syndrome (ACS). The first Thrombolysis in Myocardial Infarction (TIMI) risk score, proposed in 2000, included 7 predictor variables with one having 5 components ^1^; the HEART risk score, proposed in 2008, included 5 predictor variables with one having 6 components ^2^; the Global Registry of Acute Coronary Events (GRACE) score, proposed in 2003, included 8∼9 predictor variables ^3, 4^; the RISK-PCI risk score, proposed in 2013, included 12 predictor variables ^5^. The above-mentioned risk scores were developed based on sound statistical methods, subsequently validated in different populations, and found to yield an acceptable discrimination and calibration. Of these risk scores, the GRACE score offers the best discrimination performance and the ESC guideline recommends it be used for the assessment of all-cause mortality or non-fatal myocardial infarction in patients with acute coronary syndrome ^6^. All of these risk scores mentioned above share 2 characteristics. First, they are all designed to fit acute coronary syndrome; second, they include a number of predictor variables, where some are subject to certain degree of personal interpretation, such as prior myocardial infarction and ST-segment deviation in the TIMI risk score ^1^, family history of coronary artery disease and obesity, and diagnosed hypertension and hypercholesterolemia in the HEART risk score ^2^, Killip class, ST-segment deviation, and history of renal dysfunction in the GRACE risk score ^3, 4^, infarct-related artery reference diameter, and prior myocardial infarction in the RISK-PCI risk score ^5^. The number of predictor variables, personal interpretations of some variables, and the performance of a model will certainly limit its use in clinical practice. Therefore, if a simple model can predict the outcome with the same level of accuracy as complex models, this model should be preferred at least until a better-performing complex model appears according to the law of parsimony.

The experimental hypothesis for the present study is that a simple mortality risk score for patients with acute myocardial infarction (AMI) undergoing percutaneous coronary intervention (PCI) can be developed and may perform with adequate discrimination and calibration properties, even if the predictor variables included are minimized.

## Methods

### Study population

The CatLet extended validation trial (ChiCTR2000033730) and the CatLet validation trial (ChiCTR-POC-17013536), both being registered with chictr.org, served as the derivation and validation datasets, respectively. All the participants, aged≥18 years, homogeneous in Chinese Han ethnicity, were consecutively enrolled from the First Affiliated Hospital of Soochow University. The derivation dataset included 1018 patients admitted between January 1, 2012 and September 30, 2015, and the validation dataset included 308 ones admitted between January 1, 2012 and July 31, 2013. Both studies almost had the same baseline characteristics. Their differences are in that participants from the derivation dataset presented>12 hours since symptom onset while those from the validation dataset presented ≤12 hours. They all suffered from AMI, underwent angiography exam, and received PCI on the diseased coronary arteries. AMI diagnosis was based on the fourth universal definition of myocardial infarction ^7^. The PCI procedure was performed using the standard technique. Treatment of the diseased coronary arteries was at the discretion of the interventional cardiologists during the hospital stay. Drug-eluting stents were implanted. 300mg loading doses of aspirin and clopidogrel were prescribed before the PCI procedure; after the procedure, aspirin, 100mg/day, was prescribed lifelong, and clopidogrel, 75mg/day, was prescribed for at least one year unless contraindicated. Exclusion criteria included: (a) poor images; (b) coronary embolism; (c) coronary anomalies; and (d) normal or <50% diameter stenosis by coronary angiography. Both studies complied with the Declaration of Helsinki regarding investigation in humans and was approved by the institute review board of Soochow University. Written informed consent was obtained from all study participants or their family members.

The follow-ups were described elsewhere in detail ^8^. All-cause mortality was defined according to the recommendation by Academic Research Consortium ^9^. The scheduled follow-ups were performed by standard practice. Telephone contacts were conducted for all living patients or their immediate relatives at 1 month±7 days, 3 months±14 days, and 6 months±14 days after discharge; and thereafter, phone interviews were conducted every year±1 month. Patients were followed up for four years. Death information was obtained from Household registration management system, hospitals, or the next of kin; if conflicting death information existed, the death cause provided by hospitals was used. The follow-up was 97% completed for the derivation dataset, and was 100% completed for the validation dataset.

Information on demographic data, smoking, alcohol intake, clinical comorbidities, biochemical examinations, and lipid profiles was systematically reviewed and collected. The first admission data were collected for a patient with multiple hospitalizations. The lab results at admission were usually collected for a patient with multiple lab examinations, where the lowest value for the creatinine and the peak value for the cardiac troponin I were used for this analysis. The most recent left ventricular ejection fraction (LVEF), taken by transthoracic echocardiography within three months after discharge, was used for analysis. The 26 variables tested for association with all-cause death were as follows: age, sex, body mass index (weights in Kg/Height in meters squared), presence or absence of ST elevation myocardial infarction, primary hypertension, type 2 diabetes mellitus, stroke, smoking, alcohol intake, low-density lipoprotein cholesterol (LDL-C), high density lipoprotein cholesterol (HDL-C), triglycerides, total cholesterol, serum albumin, lipoprotein(a), cardiac troponin I, serum creatinine, fasting glucose, LVEF, monocyte, lymphocyte, white blood cell count, red blood cell count, platelet count, and hemoglobin. Their definitions and inclusion rationales were given in **online supplementary materials Table S1**. A Bonferroni correction was used to avoid type I errors caused by multiple comparisons.

### Statistical analysis and models’ development and validation

Cross-validation (10-fold) LASSO regression, a penalized multivariable logistic regression technique with a shrinkage parameter ***λ*** (lambda), was chosen to enable a rigid variable selection and less likely to overestimate the predicting value in the presence of heteroskedastic, non-Gaussian, and cluster-dependent errors. The LASSO regression places a high priority on controlling overfitting, thus often producing parsimonious models. The priority of covariate selection in a model will be given to those at a larger ***λ*** value until an added covariate does not significantly contribute to the model anymore after the ***λ*** value as shown in **online supplementary materials Table S2** ^10^. This resulted in a final parsimonious model with an acronym of BACEF: serum al**<u>B</u>**umin, **<u>A</u>**ge, serum **<u>C</u>**reatinine, and LV**<u>EF</u>**. Multicollinearity diagnostics with tolerance statistics excluded the multicollinearity among these four risk factors. Points were assigned to these four risk factors according to their respective estimated coefficients from the derivation dataset. Cutoffs of a continuous variable are selected considering two aspects: these cutoffs will yield a closest area-under-curve (AUC) to the one harvested with the continuous variable *per se*, and their clinical conveniences in use.

### Model discrimination, validation, and calibration

The model was internally validated using the K-fold cross-validation procedure. For internal-external (temporal) validation, the derivation dataset were non-randomly split ^11^. The same model was developed in the earlier two-thirds (n=679) and validated in the latter third (n=339) of the derivation population. Finally, external validation was performed in 308 patients from the CatLet validation trial.

Discrimination was assessed by the AUC of receiver operating characteristic analysis (ROC). Calibration was evaluated using the intercept and slope showing the agreement between the observed and predicted mortality risk. Clinical usefulness was assessed with decision curve analysis. Kaplan-Meier curves were used to show the mortality stratified by the tertile of the BACE score. It was also compared with ACEF score ^12^, ACEFII score ^13^, GRACE score ^4^ with respect to discrimination by the net reclassification index (NRI) and integrated discrimination index (IDI). A cutoff equal to the event rate would be used for binary classification, and cutoffs equal to half the event rate, the event rate, and twice the event rate would be used when more than 2 categories are desired ^14^. In the current study, the average mortality rate was 15% (198/1326) for both derivation and validation datasets. Thus, half the event rate, the event rate, and twice the event rate are 7.5%, 15%, and 30%, respectively. These three cutoffs categorized the external validation cohort in this study: <7.5%, 7.5-15%, 15-30%, and >30%.

Continuous variables were compared using the Mann-Whitney *U* test and categorical variables were compared using Pearson’s Chi-squared test. All the calculations and graphs were performed using STATA 16 (State Corp LP, College Station, TX, USA). The TRIPOD Statement was applied for derivation, validation, and reporting of risk prediction models ^11^. All tests were two-sided. A value of *P* < 0.05 was considered significant for all statistical tests.

## Results

**Table 1** reported the baseline characteristics of the score’s development cohort (CatLet extended validation trial, n=1018) and the external validation cohort (CatLet validation trial, n=308). Patients’ inclusion and exclusion has been detailed in online supplementary Figure S1. Over the four years of follow-ups, the validation cohort had a higher death rate (55 (17.86%) vs 143 (14.05%)) as compared with derivation cohort.

**Table 1.**
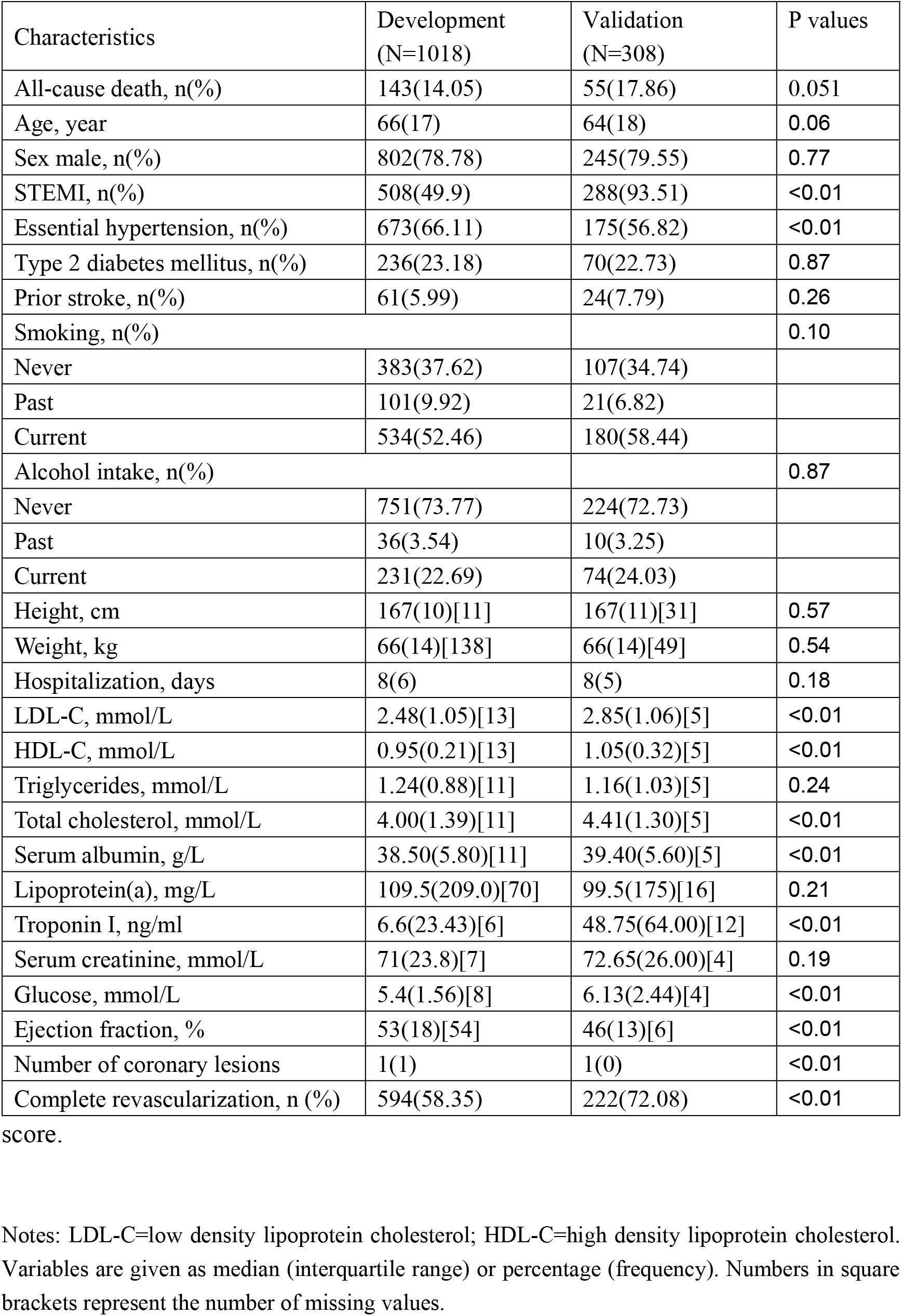
Baseline characteristics for development and external validation of the BACEF

| Characteristics | Development<br>(N=1018) | Validation<br>(N=308) | P values |
| --- | --- | --- | --- |
| All-cause death, n(%) | 143(14.05) | 55(17.86) | 0.051 |
| Age, year | 66(17) | 64(18) | 0.06 |
| Sex male, n(%) | 802(78.78) | 245(79.55) | 0.77 |
| STEMI, n(%) | 508(49.9) | 288(93.51) | <0.01 |
| Essential hypertension, n(%) | 673(66.11) | 175(56.82) | <0.01 |
| Type 2 diabetes mellitus, n(%) | 236(23.18) | 70(22.73) | 0.87 |
| Prior stroke, n(%) | 61(5.99) | 24(7.79) | 0.26 |
| Smoking, n(%) |  |  | 0.10 |
| Never | 383(37.62) | 107(34.74) |  |
| Past | 101(9.92) | 21(6.82) |  |
| Current | 534(52.46) | 180(58.44) |  |
| Alcohol intake, n(%) |  |  | 0.87 |
| Never | 751(73.77) | 224(72.73) |  |
| Past | 36(3.54) | 10(3.25) |  |
| Current | 231(22.69) | 74(24.03) |  |
| Height, cm | 167(10)[11] | 167(11)[31] | 0.57 |
| Weight, kg | 66(14)[138] | 66(14)[49] | 0.54 |
| Hospitalization, days | 8(6) | 8(5) | 0.18 |
| LDL-C, mmol/L | 2.48(1.05)[13] | 2.85(1.06)[5] | <0.01 |
| HDL-C, mmol/L | 0.95(0.21)[13] | 1.05(0.32)[5] | <0.01 |
| Triglycerides, mmol/L | 1.24(0.88)[11] | 1.16(1.03)[5] | 0.24 |
| Total cholesterol, mmol/L | 4.00(1.39)[11] | 4.41(1.30)[5] | <0.01 |
| Serum albumin, g/L | 38.50(5.80)[11] | 39.40(5.60)[5] | <0.01 |
| Lipoprotein(a), mg/L | 109.5(209.0)[70] | 99.5(175)[16] | 0.21 |
| Troponin I, ng/ml | 6.6(23.43)[6] | 48.75(64.00)[12] | <0.01 |
| Serum creatinine, mmol/L | 71(23.8)[7] | 72.65(26.00)[4] | 0.19 |
| Glucose, mmol/L | 5.4(1.56)[8] | 6.13(2.44)[4] | <0.01 |
| Ejection fraction, % | 53(18)[54] | 46(13)[6] | <0.01 |
| Number of coronary lesions | 1(1) | 1(0) | <0.01 |
| Complete revascularization, n (%) | 594(58.35) | 222(72.08) | <0.01 |

score.
Notes: LDL-C=low density lipoprotein cholesterol; HDL-C=high density lipoprotein cholesterol. Variables are given as median (interquartile range) or percentage (frequency). Numbers in square brackets represent the number of missing values.

### Development of the BACEF score and its performance and internal validation

By applying the procedure specified before, the penalized multivariable logistic regression technique revealed four risk factors, namely serum albumin, age, serum creatinine, and LVEF to predict 4-year mortality. These four risk factors, all predictive of all-cause mortality in univariate analysis as shown in **in online materials Table S3**, were subsequently entered into a multivariable logistic regression model and confirmed to be predictive of all-cause mortality as shown in **Table 2**. Points were assigned to these four risk factors according to their respective estimated coefficients from the derivation dataset as shown in **Table 3 and in online materials Table S4**.

**Table 2.** Multivariable Logistic regression analysis for the associations of the four risk factors with mortality risk.

| Risk factors | Coefficients | t | P-values | OR(95%CI) |
| --- | --- | --- | --- | --- |
| Serum albumin, g/L | -0.1090607 | -4.75 | P<0.001 | 0.90(0.86-0.94) |
| Age, years | 0.0737409 | 6.43 | P<0.001 | 1.08(1.05-1.10) |
| Serum creatinine, mg/dL | 0.6771199 | 4.23 | P<0.001 | 1.97(1.44-2.69) |
| LVEF, % | -0.0591556 | -5.79 | P<0.001 | 0.94(0.92-0.96) |
| Constants | -0.6266035 | 0.639 | 0.731 | 0.53(0.04-7.30) |

Multivariable diagnostics: albumin, 0.830 tolerance; age, 0.844 tolerance; EF, 0.923 tolerance; serum creatinine, 0.970. ORs per 1 unit higher.

**Table 3.** Point assignment to the four predictor variables.

| Risk Factor | Categories | Points |
| --- | --- | --- |
| Serum albumin, g/L | $\geq 40$ | 0 |
| | $35 \leq \text{albumin} < 40$ | 1 |
| | $30 \leq \text{albumin} < 35$ | 2 |
| | $< 30$ | 3 |
| Age, years | $< 45$ | 0 |
| | $45 \leq \text{age} < 65$ | 1 |
| | $65 \leq \text{age} < 75$ | 2 |
| | $75 \leq \text{age} < 80$ | 3 |
| | $\geq 80$ | 4 |
| Ejection fraction, % | $\geq 60$ | 0 |
| | $50 \leq \text{LvEF} < 60$ | 1 |
| | $40 \leq \text{LvEF} < 50$ | 2 |
| | $< 40$ | 3 |
| Serum creatinine, mg/dL | $< 1.0$ | 0 |
| | $1.0 \leq \text{Creatinine} < 1.5$ | 1 |
| | $\geq 1.5$ | 2 |

**FIGURE 1** showed that BACEF score significantly predicted the mortality risk for patients with AMI. The discrimination was good with an AUC (95%CI) of 0.84(0.80-0.87). The 10-fold cross-validation revealed an AUC of 0.83, similar to the one obtained from the full dataset. The calibration plot and the decision curve analysis for the BACEF score in the whole derivation dataset were presented in **FIGURE S2 and FIGURE S3, respectively**.

**Figure 1.**
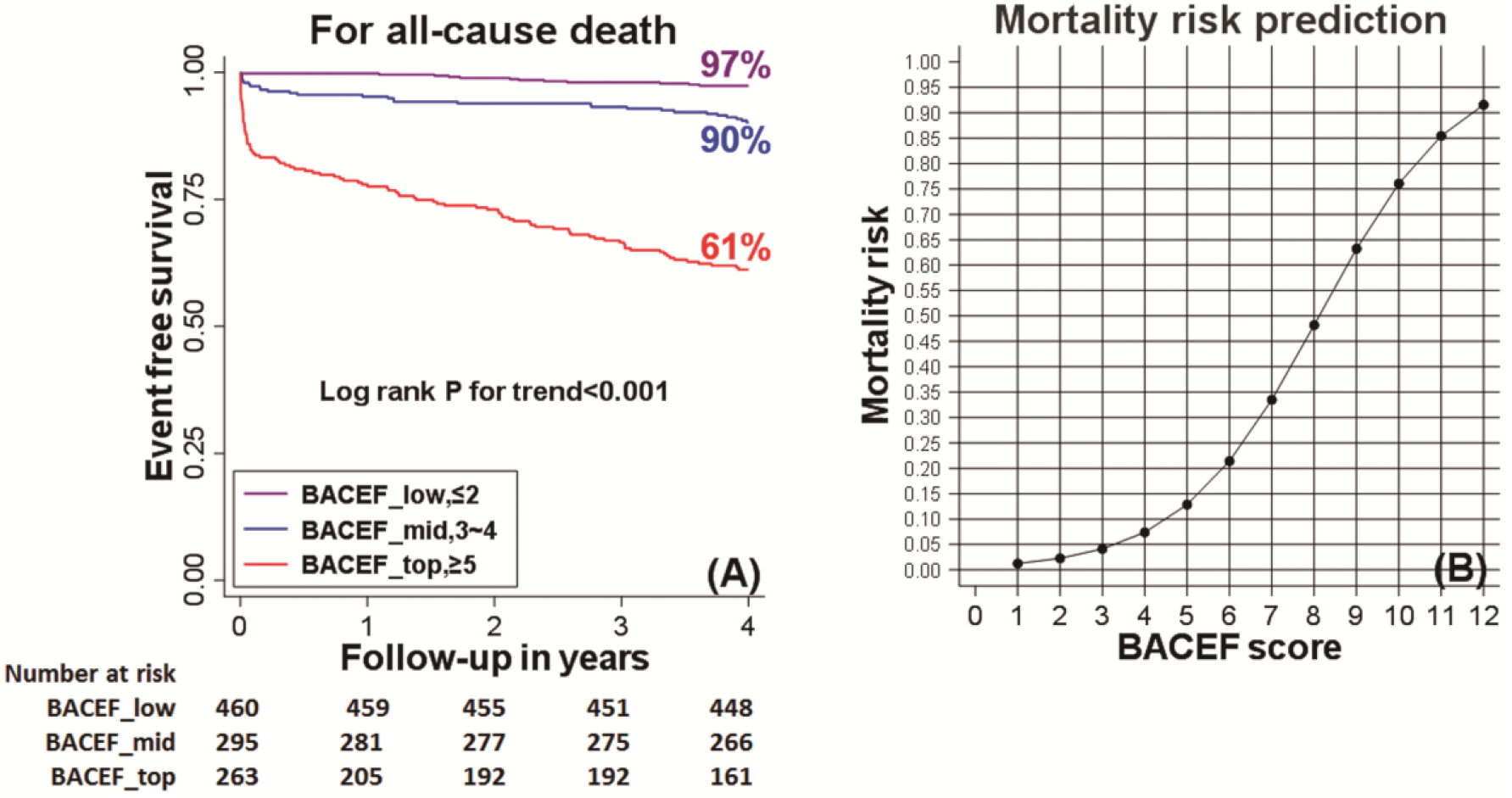
BACEF score and its univariate associations according to the Kaplan-Meier survivor function (Figure 1A) and the Logistic regression analysis (Figure 1B), respectively.

### Internal-external (temporal) validation of the BACEF score

The same predictive models based on the four clinical variables (BACEF) were developed in the earlier two-thirds (n=679) of the derivation dataset for temporal validation. In the later third of the validation dataset (n=339), they yielded an AUC statistic of 0.89(0.83-0.95). The BACEF score was well calibrated and showed a large positive net benefit according to the decision curve analysis in the internal-external validation cohort as shown in **Figure S4 and Figure S5**.

### External validation of the BACEF score

The BACEF score was externally validated in 308 patients from the CatLet validation trial ^15^. In terms of discrimination, the BACEF score yielded the AUC of 0.83(95% CI (0.77-0.89)). The calibration and recalibration plots and the decision curve analysis in the external validation cohort were all good as displayed in **Figure S6A-S6B and Figure S7**.

### Comparison with other clinically-based scoring systems

In the external validation cohort, **Figure 2** showed the best performance of the BACEF score (AUC (95%CI), 0.83(0.77-0.89)) and the poorest performance of the GRACE score (0.79(0.72-0.86)). Better performance with the BACEF score has been confirmed by the net reclassification index (NRI) or integrated discrimination index (IDI) as compared with the other three already existent risk-predicting scores as shown in **TABLE 4**. Calibration was good with BACEF score (P>0.05) and poor with ACEF, ACEF II, and GRACE score (all P<0.05).

**Figure 2.**
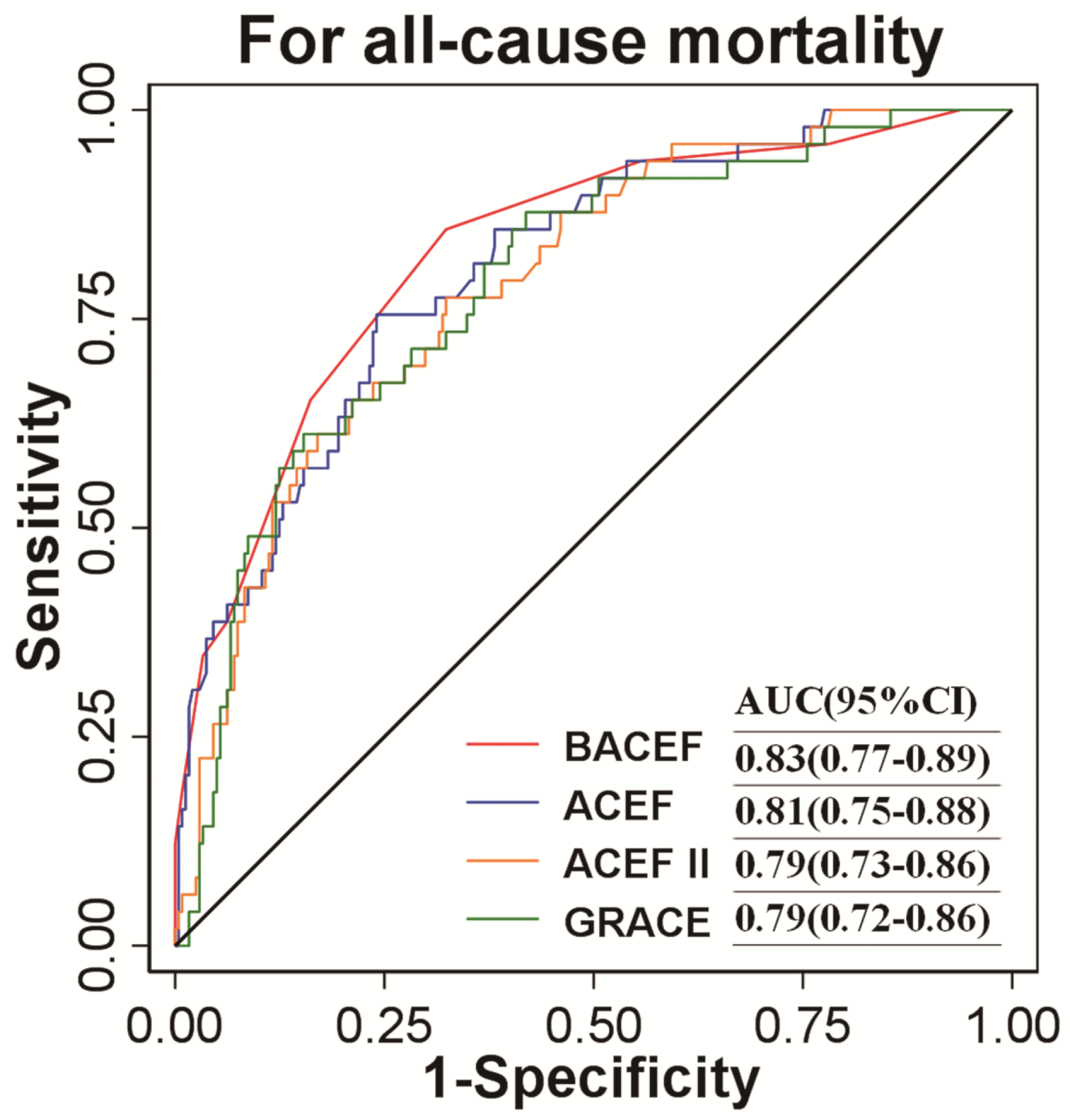
ROC curves for the four risk scores in the external validation cohort.

**Table 4.** The net reclassification index (NRI) and the integrated discrimination index (IDI) of the risk-predicting models as compared with the BACEF score in the external validation cohort.

|  | ACEF | ACEF II | GRACE |
| --- | --- | --- | --- |
| Participants, n | 308 | 308 | 308 |
| Death events, n | 55 | 55 | 55 |
| Category-dependent NRI | 0.28 | 0.63 | 0.49 |
| *P* values | <0.05 | <0.05 | <0.05 |
| Category-free NRI | 0.35 | 0.77 | 0.80 |
| *P* values | <0.05 | <0.05 | <0.05 |
| IDI | 0.05 | 0.13 | 0.12 |
| *P* values | 0.05 | <0.05 | <0.05 |

Risk categories: <7.5%、7.5-15%、15-30% 及 >30%

## Discussion

The key findings in the current study are as follows: i) a simple risk score with an acronym of BACEF, including four risk factors of serum albumin, age, serum creatinine, and LVEF, has been developed, predicting the 4-year mortality risk well in AMI patients; ii) this novel BACEF score has been extensively validated; and iii) this BACEF score has outperformed the ACEF, ACEF II, or GRACE score in terms of prognostication in the external validation cohort.

### The BACEF score and its four predictor variables

A simple model only based on serum albumin, age, serum creatinine, and LVEF has claimed that it performs equivalently to or even better than the existing ACS risk scores in terms of mortality prediction. The exclusion of a large number of risk factors and comorbidities will naturally cause a concern. However, when we carefully look at the components of TIMI, GRACE, and RISK-PCI risk scores, age, serum creatinine, and LVEF are all present in these risk scores ^1, 4, 5^; and these three risk factors are also present in many risk scores for patients receiving cardiac surgery, such as ACEF, ACEF II, EuroSCORE ^12, 13, 16^. The ACEF score including only these three risk factors, originally developed to predict operative mortality for patients receiving cardiac surgery, also performed well with respect to mortality prediction for CAD patients, with the C-statistics being 0.73∼0.76 ^17, 18^. Considering the robust predictive values of these three risk factors, we may rest assured that our parsimonious BACEF model will also perform well in terms of mortality prediction for AMI patients. The concern due to its exclusion of a large number of risk factors and comorbidities seems to be mitigated. The inclusion of serum albumin into a risk score is a novelty. In 2004, we have observed that low serum albumin is predictive of the occurrence of atrial fibrillation ^19^. Our subsequent studies have further confirmed the association of the low serum albumin with the occurrence or outcomes of cardiovascular diseases (CVD) ^20-23^. A meta-analysis including 14 studies with 150 652 individuals without prior cardiovascular diseases (CVD), has shown that per 10 g/L lower serum albumin was associated with a 1.85-fold increased risk of a CVD event during a median follow-up of 8.5 years ^24^.

Another Meta-analysis included 8 studies with 21,667 ACS patients, showing that hypoalbuminemia had an increased risk of all-cause mortality (2.15(95% CI, 1.68-2.75)) after adjusting for covariates ^25^. There is now ample evidence that serum albumin, a marker of easy accessibility and definite objectivity, is a powerful, unrecognized prognostic marker in the general population as well as in many disease states. In the current study, serum albumin has the second best AUC statistic, only secondary to the age in the univariate analysis. The physiological actions of the serum albumin include its anti-platelet aggregation, anti-coagulant properties, anti-oxidative activity, and anti-inflammatory activity, which can explain the association of low serum albumin levels with the adverse cardiovascular outcomes ^26^.

### The performance of the BACEF score and its comparison with three other risk scores

In the derivation cohort, the BACEF score has performed well in terms of discrimination and calibration. In the external validation cohort, the BACEF score had the best performance while the GRACE score had the poorest performance in terms of mortality prediction. The ACEF score had the second best performance with respect to mortality prediction for AMI patients in the current study ^12^. The excellent performance of the ACEF score in the other coronary artery disease populations has been widely validated ^17, 18^. In the external validation cohort, as compared with the ACEF score having the second best performance, the BACEF score also proved a significantly better discrimination confirmed by the NRI and the IDI, and was also better calibrated (***χ***^**2**^=10.47, ***P***=0.234). The strengths of the BACEF score include that categorization of all the four continuous variables has eased its clinical use; a better discrimination and calibration with the BACEF score has been achieved in the context of categorized predictors as compared with the ACEF score, in which, age and LVEF were treated as continuous variables; serum albumin is accessible and is definitely not subject to personal estimation; and the BACEF score still do not betray the law of parsimony. These strengths have supported the use of the BACEF score as a risk-stratifying tool for AMI patients.

### Definitions of predictor variables

The four predictor variables included in the BACEF score are all continuous variables. Three of them (serum albumin, age, and serum creatinine) are definitely not subject to personal estimation. The LVEF could be less defined because of patients’ different situations or different echo cardiologists. In our tertiary medical center, echocardiography evaluations were routinely conducted at the end of three months after discharge. We chose to utilize this value for analysis as the LVEF values for AMI patients usually level off after three months later. This provides a relatively standardized assessment of this predictor variable. The BACEF score does not include categorical predictor variables, such as diabetes, smoking, or hypertension. Introduction of these variables will be subject to certain degree of personal interpretation although they are usually clearly defined in each risk score. Interestingly, the priority of covariate selection provided by the LASSO regression supports the exclusion of any categorical predictors in the current study. Chronic obstructive pulmonary disease is a common comorbidity incorporated into surgery-related risk scores such as the EuroSCORE ^16^. Unfortunately, we did not collect the information on this comorbidity in the current study. The ACEF score included only three risk factors of age, serum creatinine, and LVEF, without chronic obstructive pulmonary disease, and outperformed most surgery-related risk scores in terms of outcome predictions on one hand ^12^, on the other hand, most currently available ACS risk scores did not include chronic obstructive pulmonary disease ^1-5^. These seem to justify the BACEF score’s exclusion of chronic obstructive pulmonary disease.

### Study limitations

The current study enrolls only AMI patients; therefore, the BACEF score cannot be applied to the whole CAD population. Second, this novel risk score is not validated in different hospitals or in different patient populations. When a risk score is applied in a different population, we can use the risk score directly, or recalibrate the model via adjusting the slope and the intercept; we also can refit the new different population using the four predictor variables ^27^. Finally, we may miss out new risk factors that result in a better model performance for outcome predictions for ACS patients using the less, the same, or the more number of risk factors although we have conducted a thorough literature search for a possible risk factor that may have an incremental value for the model.

## Conclusion

This novel BACEF risk score, including only four predictor variables (serum albumin, age, serum creatinine, and LVEF) presents itself as a parsimonious and easy-to-use tool for mortality risk stratification in AMI patients, waiting for adequately sized external populations to undergo the imperative validation process.

## Data Availability

All data produced in the present study are available upon reasonable request to the authors

## Abbreviations

Abbreviations: Full names
ACEF: Age, serum creatinine, and LVEF
ACS: Acute coronary syndrome
AMI: Acute myocardial infarction
AUC: Area-under-curve
BACEF: Serum albumin, age, serum creatinine, and LVEF
CatLet: Coronary Artery Tree description and Lesion EvaluaTion
GRACE: Global Registry of Acute Coronary Events
LVEF: Left ventricular ejection fraction
PCI: Percutaneous coronary intervention
ROC: Receiver operating characteristic
TIMI: Thrombolysis in Myocardial Infarction

## Acknowledgement

None.

## Funding

This work was in part supported by the National Key R&D Program (2020YFC2004705), Sci-Tech Supporting Program of Jiangsu Commission of Health (M2021019), and Medical Sci-Tech innovation Program for Medical Care of Suzhou City (SKY2021005).

## Conflict of interest

None declared.

## Authors’ contribution

He YM drafted and refined the manuscript. He YM, Tao JY, and Ge ZY conceived and designed the work. He YM and Ge ZY analyzed and interpreted the data for this work. Jiang TB and He Y collected the data for this work. All gave final approval and agree to be accountable for all aspects of work ensuring integrity and accuracy.

## Data available statement

Individual participant data are available when appropriate.

## Notes

### Competing Interest Statement

The authors have declared no competing interest.

### Author Declarations

Both studies complied with the Declaration of Helsinki regarding investigation in humans and was approved by the institute review board of Soochow University. Written informed consent was obtained from all study participants

